# Using computational approaches to enhance the interpretation of missense variants in the *PAX6* gene

**DOI:** 10.1101/2023.12.21.23300370

**Authors:** Nadya S. Andhika, Susmito Biswas, Claire Hardcastle, David Green, Simon C. Ramsden, Ewan Birney, Graeme C. Black, Panagiotis I. Sergouniotis

## Abstract

**Purpose:** The *PAX6* gene encodes a highly-conserved transcription factor involved in eye development. Heterozygous loss-of-function variants in *PAX6* can cause a range of ophthalmic disorders including aniridia. A key molecular diagnostic challenge is that many *PAX6* missense changes are presently classified as variants of uncertain significance. While computational tools can be used to assess the effect of genetic alterations, the accuracy of their predictions varies. Here, we evaluated and optimised the performance of computational prediction tools in relation to *PAX6* missense variants.

**Methods:** Through inspection of publicly available resources (including HGMD, ClinVar, LOVD and gnomAD), we identified 241 *PAX6* missense variants that were used for model training and evaluation. The performance of ten commonly-used computational tools was assessed and a threshold optimization approach was utilized to determine optimal cut-off values. Validation studies were subsequently undertaken using *PAX6* variants from a local database.

**Results:** AlphaMissense, SIFT4G and REVEL emerged as the best-performing predictors; the optimized thresholds of these tools were 0.967, 0.025, and 0.772, respectively. Combining the prediction from these top-three tools resulted in lower performance compared to using AlphaMissense alone.

**Conclusion:** Tailoring the use of computational tools by employing optimized thresholds specific to *PAX6* can enhance algorithmic performance. Our findings have implications for *PAX6* variant interpretation in clinical settings.

## INTRODUCTION

The *PAX6* gene (Paired box 6, OMIM #607108, HGNC 8620) encodes a DNA-binding protein that performs essential regulatory functions during eye development in many animal species including humans.^1,2^ Genetic variants in *PAX6* underlie a number of ophthalmic disorders. By far the most common *PAX6-*related oculopathy is aniridia (OMIM #106210), a condition associated with *PAX6* haploinsufficiency due to heterozygous loss-of-function variants.^3^ Missense variants have been generally linked with milder phenotypes.^4,5^ However, in 2020, a study by Williamson *et al*. highlighted that certain heterozygous *PAX6* missense variants can cause clinical manifestations that are more severe than aniridia (including microphthalmia and anophthalmia).^6^ Predicting the effect of the growing number of missense variants that are being identified remains challenging. Notably, when established criteria (such as those described by the American College of Medical Genetics and Association of Molecular Pathology (ACMG/AMP)) are used to classify these sequence alterations, a significant proportion are classified as variants of uncertain significance (VUS).^7,8^

Computational *(in silico)* tools are commonly used to provide evidence to support or refute variant pathogenicity.^8^ Each tool employs a different algorithm; features commonly taken into account include evolutionary conservation and protein/domain structure (**Supplementary Table 1**). It is noted that some algorithms combine the output from other tools to achieve a single consensus prediction (meta-predictors).^9^

A number of previous studies have evaluated the performance of commonly-used computational tools in different genes, noting significant variability in predictive performance.^10–13^ Aiming to increase the reliability of existing algorithms and to optimise their predictions, some studies have proposed the introduction of gene-specific thresholds.^14,15^ To date, computational tool evaluation and optimization have not been undertaken in the context of *PAX6* and this study aims to address this gap.

## MATERIALS AND METHODS

### Dataset collection

In our primary analysis, *PAX6* missense variants from publicly available resources were collected from: the Genome Aggregation Database (gnomAD) version 2.1.1 (v2) and version 3.1.1 (v3); the Leiden Open Variation Database (LOVD) version 2.0 and version 3.0; the Human Genetic Mutation Database (HGMD) Public version; and ClinVar (the websites of these resources can be found in the ‘Web Resources’ section) (all accessed in February 2023). A biomedical literature search (MEDLINE/PubMed) using the term "PAX6" and focusing on articles between 2021 and 2023 was also undertaken.^16–20^ We excluded duplicates and VUS (including “likely disease-causing mutation with questionable pathogenicity” (DM?) in HGMD), and then categorised the remaining variants into: “Primary Dataset Neutral” and “Primary Dataset Disease” (**Figure 1**).

**Figure 1.**
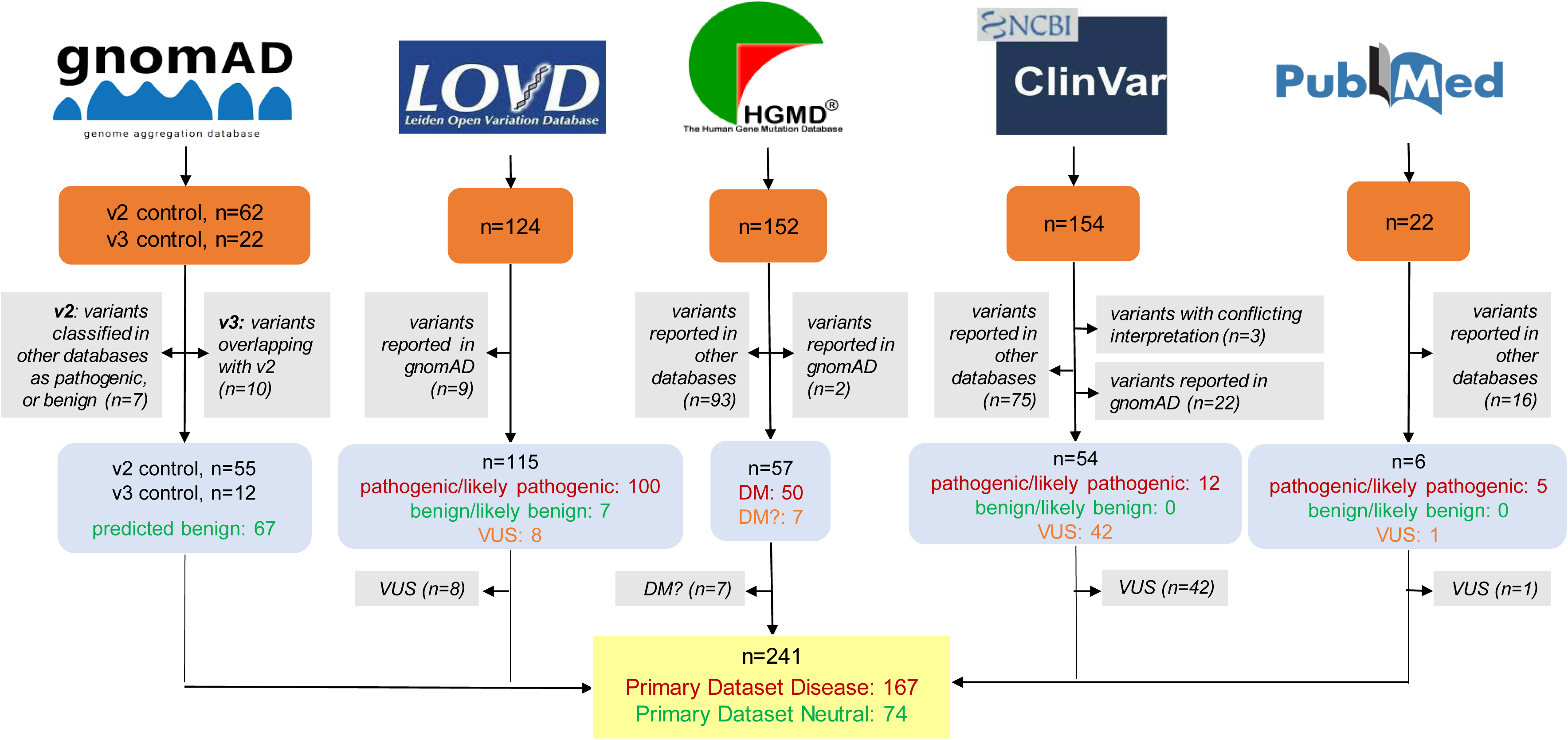
Overview of the datasets used in the primary analysis. The utilized resources, selection criteria and filtering steps are outlined. Orange boxes: number of compiled variants; grey boxes: exclusion criteria and number of excluded variants; blue boxes: number of filtered variants based on their pathogenicity; yellow box: number of variants in the two sub-datasets. gnomAD, Genome Aggregation Database; LOVD, Leiden Open Variation Database; HGMD, Human Gene Mutation Database; DM, “disease-causing mutation” (as assigned in HGMD); DM?, “likely disease-causing mutation with questionable pathogenicity” (as assigned in HGMD); VUS, variants of uncertain significance. All five resources were accessed in February 2023.

Primary Dataset Neutral included: (i) variants previously classified as benign or likely benign and (ii) variants present in gnomAD, a population-scale database that does not include individuals with severe paediatric disease.^16^ While it cannot be excluded that certain *PAX6* missense variants reported in gnomAD are pathogenic (e.g. if linked with subclinical phenotypes or incomplete penetrance), we adopted a pragmatic approach and considered these changes as “presumed benign”. Although filtering gnomAD variants based on their allele frequency would increase the likelihood of including only truly benign variants, this would reduce the dataset size. Hence, we did not apply such a filter. Primary Dataset Disease included missense variants labelled as pathogenic in ClinVar, LOVD or PubMed and variants labelled as DM in HGMD.

For validation purposes, a secondary analysis was conducted involving *PAX6* missense variants from our local database at the Manchester Centre for Genomic Medicine (MCGM), part of the North West Genomic Laboratory Hub (accessed in May 2023). These variants correspond to changes that were evaluated in an accredited diagnostics laboratory with >15 years’ experience in assessing genetic alterations from individuals with ophthalmic disorders. All variants were classified according to the ACMG/AMP 2015 guidelines^8^ and changes assigned to the “likely pathogenic” and “pathogenic” categories formed the “Secondary Dataset Disease” (**Figure 2**). For this replication study, variants present in the BRAVO database (version TOPMed Freeze 8) were collected (accessed in May 2023) and formed “Secondary Dataset Neutral”. Duplicates were excluded, while the detected VUS were used for downstream analysis.^21^

**Figure 2.**
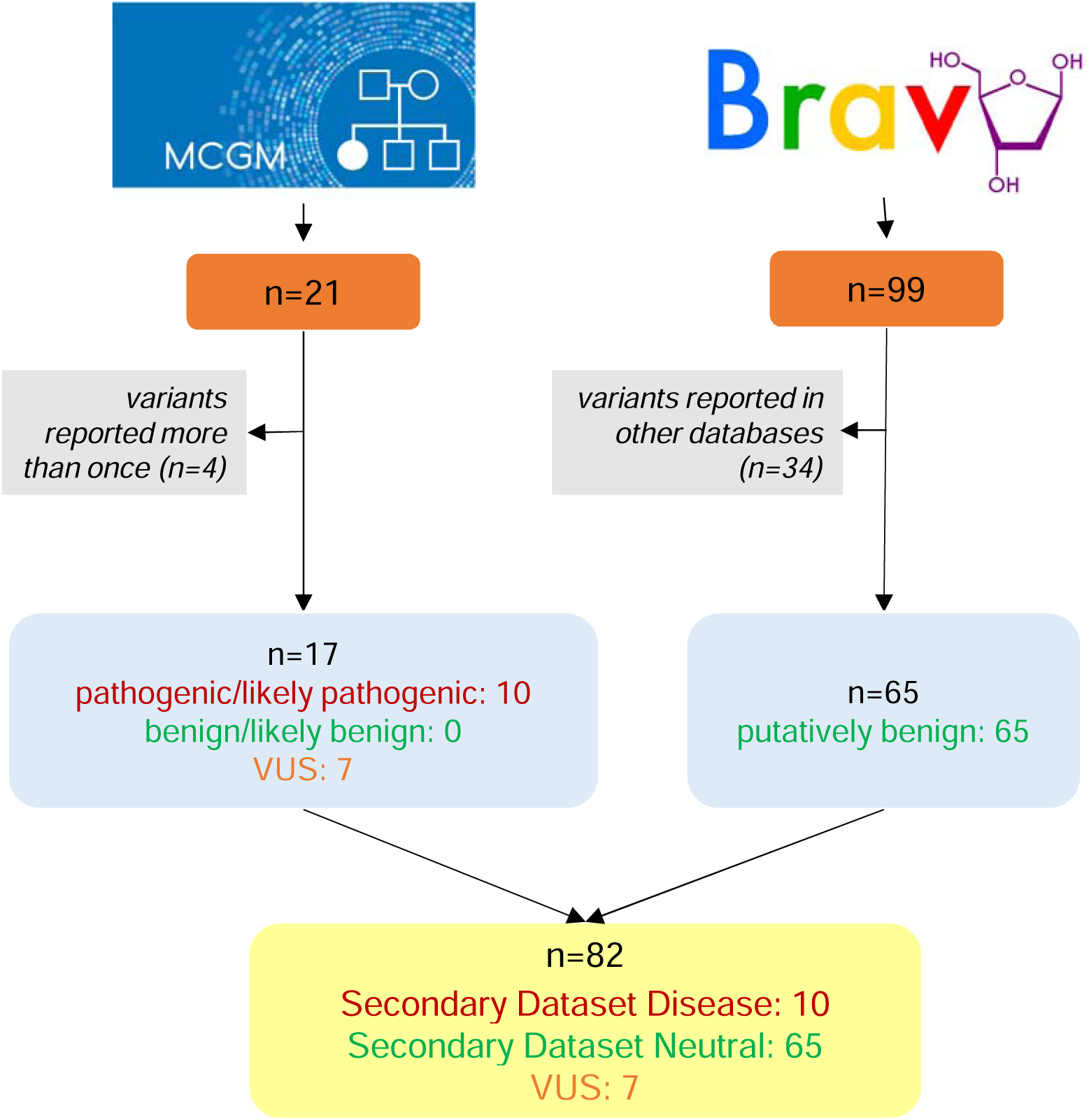
Overview of the datasets used in the secondary analysis. The utilized resources, selection criteria and filtering steps are outlined. Orange box, number of compiled variants; grey box, exclusion criteria and number of excluded variants; blue box, number of filtered and compiled variants based on their pathogenicity; yellow box, number of variants grouped into two sub-datasets and the number of VUS collected. MCGM, Manchester Centre for Genomic Medicine; VUS, Variant of Uncertain Significance. Both resources were accessed in May 2023.

All variants were numbered based on Genome Reference Consortium Human Build 38 (GRCh38). Variants from gnomAD v2 were lifted over to this reference, using the transcript ENST00000241001 (Ensembl ID), which encodes the canonical PAX6 protein, comprising 422 amino acids (UniProt ID: P26367-1).^22^

### Descriptive analysis

The distribution of variants in Primary Dataset Disease, Primary Dataset Neutral, Secondary Dataset Disease and Secondary Dataset Neutral along the linear protein sequence (as retrieved from UniProt) was visualised using a lolliplot diagram. The cBioPortal (version 5.4.5) tool was used to generate the relevant figure (accessed in May 2023) (**Figure 3**).^23^

**Figure 3.**
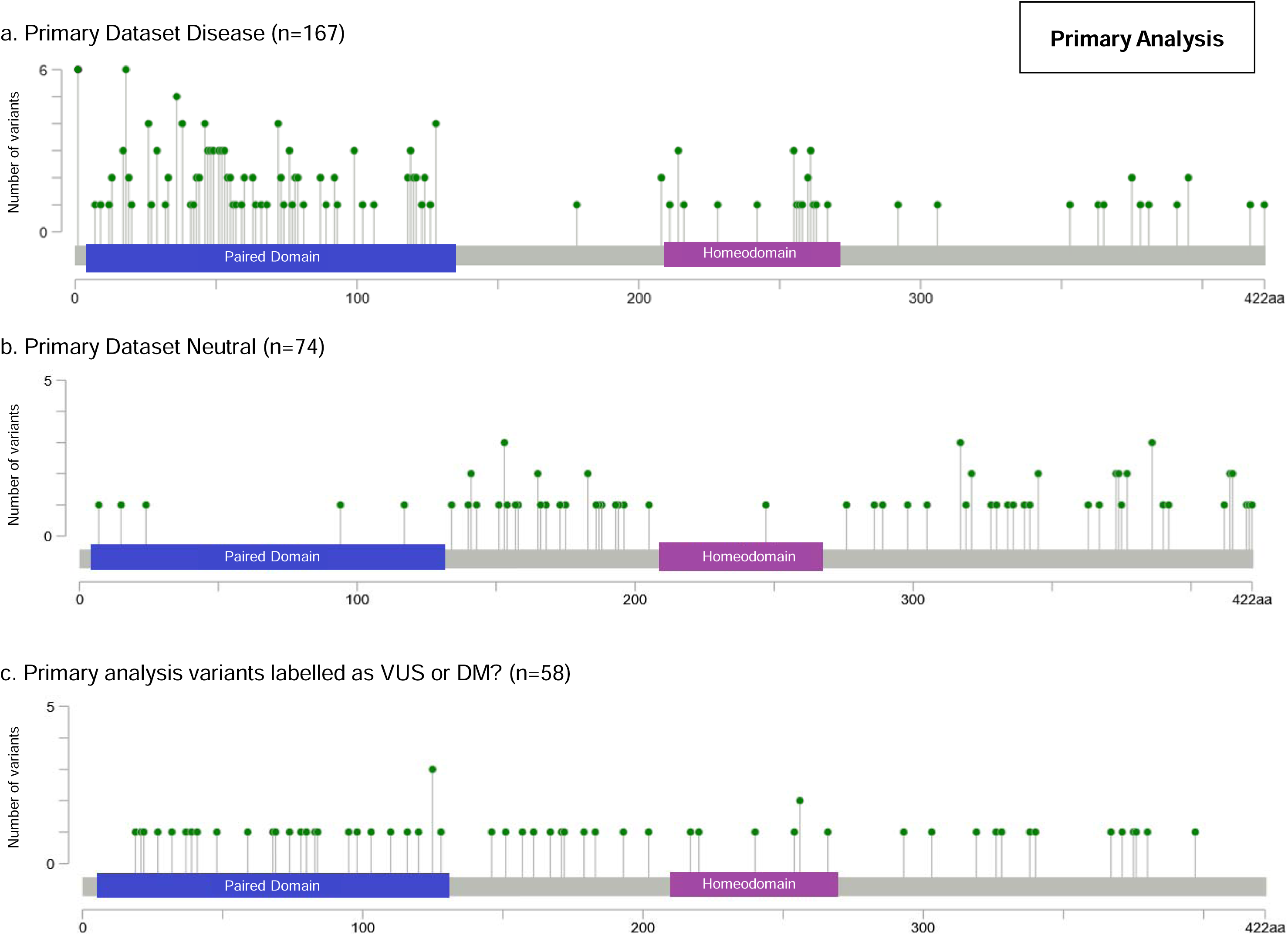
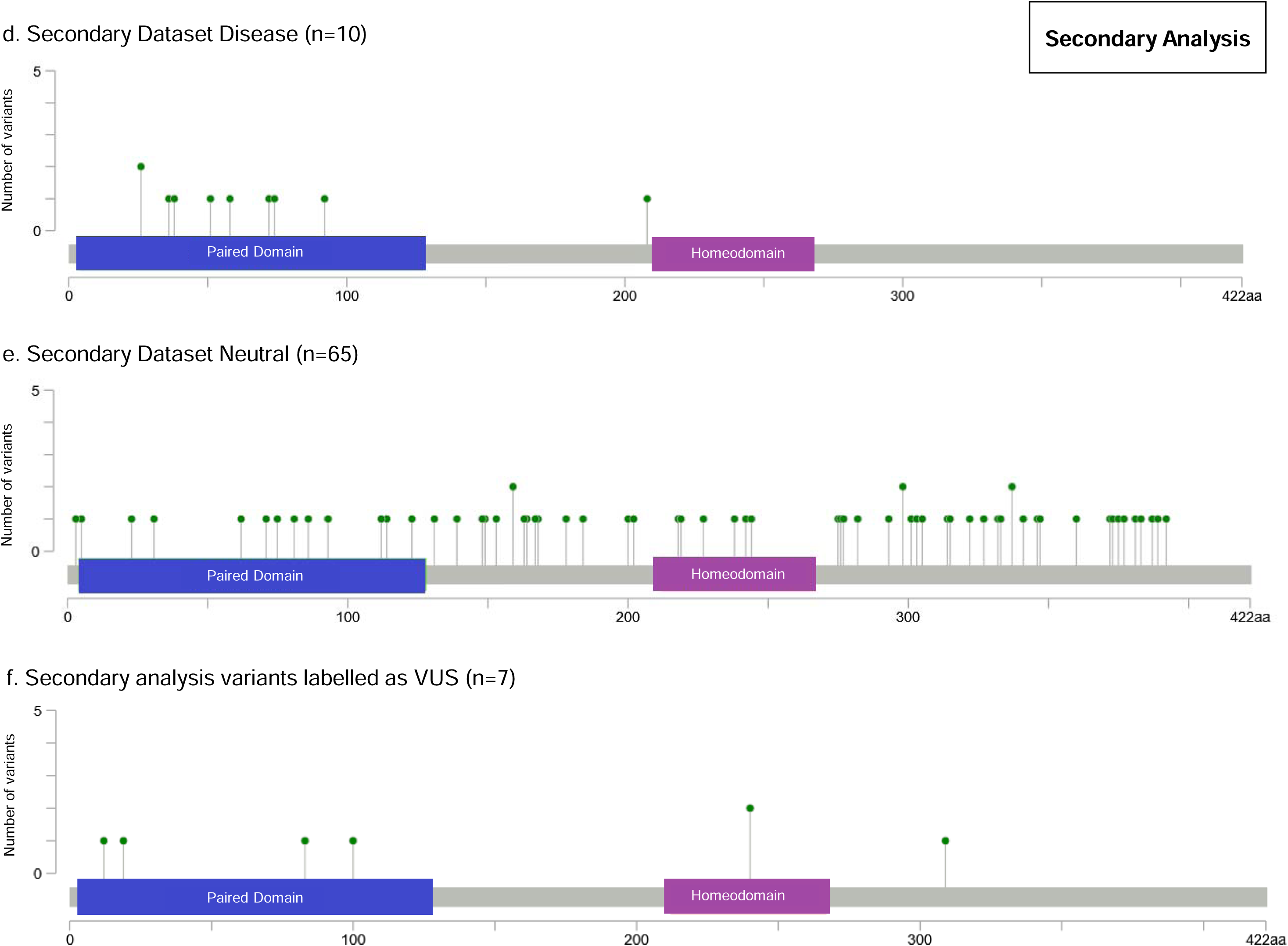
Distribution of the *PAX6* missense variants included in this study. Variant distribution according to their pathogenicity is shown. The X-axis represents the PAX6 canonical protein sequence (422 amino acids), while the Y-axis denotes the number of variants impacting the same residue. Blue bar, paired domain; purple bar, homeodomain. DM?, “likely disease-causing mutation with questionable pathogenicity” (as assigned in the Human Gene Mutation Database [HGMD]); VUS, variants of uncertain significance.

### Computational tools

Ten commonly used computational prediction tools were assessed: AlphaMissense, BayesDel, CADD, ClinPred, Eigen, MutPred2, Polyphen-2, REVEL, SIFT4G and VEST4.^24–33^ These tools employ various algorithms to evaluate variant pathogenicity (more information on the utilized approaches can be found in **Supplementary Table 1**). The dbNSFP (version 4.1) resource was used to obtain pathogenicity scores for each tested variant except for AlphaMissense. AlphaMissense prediction scores were extracted from the AlphaMissense_hg38.tsv.gz file provided in the relevant publication.^24^

Depending on how the obtained scores compared to each algorithm’s pre-set threshold (determined by the respective tool’s developers), the studied variants were classified as “predicted pathogenic” or “predicted benign”.^34^ Default thresholds were set for CADD and Eigen based on previous studies (although the use of a single, arbitrary threshold is not recommended by the tools’ developers). For AlphaMissense, we assigned variants with scores ranging from 0.564 to 1.00 to the “predicted pathogenic” category; all other variants were assigned to a “predicted benign” group. Higher scores indicated a higher likelihood of a pathogenic prediction for all tools except SIFT4G. In a few cases, a single tool generated multiple scores and we opted for the following: CADD-phred; BayesDel AddAF (incorporates allele frequency data); Eigen raw for coding variants; and the PolyPhen-2 HumVar-trained model, which is suitable for studying Mendelian diseases.^26^ The prediction outputs “deleterious”, “damaging”, “probably damaging”, or “possibly damaging” were considered “predicted pathogenic”, while the terms “tolerated” or “benign” were deemed “predicted benign”.

### Performance assessment

Initially, performance parameters were calculated using the *PAX6* missense variants included in the primary datasets. We estimated sensitivity, specificity, accuracy, precision (Positive Predictive Value; PPV), and the Matthews Correlation Coefficient (MCC).^35^ To determine the best-performing tool, we used MCC, which ranges from - 1 (constant false predictions) to 1 (perfect predictions) with 0 indicating random prediction.

We hypothesized that using an optimized, gene-specific threshold can improve the performance of each tool. Receiver Operating Characteristic (ROC) curves were therefore utilized to identify the threshold that yielded the highest MCC score for each tool. The quality of the prediction obtained using the optimized threshold was then compared to that obtained using the default threshold. The IBM SPSS (Version 25.0)^36^ software was used for these analyses.

Subsequently, we explored if the analytical performance could be further improved by combining the three tools with the highest MCC scores into a custom meta-predictor. We adopted the "majority rule" method (agreement of over 50% of the employed tools), which involved classifying a variant as "predicted pathogenic" if it received a "predicted pathogenic" score in at least two out of the three selected tools.

### Validation and evaluation

We validated our findings using a 5-fold cross-validation approach for the tool with the highest MCC score (as previously described by Tang *et al*.^11^). Briefly, this involved randomly dividing variants into five subsets of equal size, four of which (80%) formed the training set, while the remaining subset (20%) served as the test set. Within the training set, we determined the optimized threshold that maximized the MCC. The obtained threshold was then applied to assess performance on the testing set. This process was repeated five times until all subsets were utilized as the testing set. The resulting analytical pipeline was then evaluated on a secondary dataset and was used to evaluate a set of variants that were previously classified as VUS.

## RESULTS

### *PAX6* variant datasets

In our primary analysis, we collected a total of 241 variants from publicly available databases. Using pre-determined criteria (see Methods) these were split into two groups: Primary Dataset Disease (n=167) and Primary Dataset Neutral (n=74) (**Figure 1**). For the secondary analysis, we collected 17 unique variants from our local database, consisting of seven that were classed as VUS and 10 classed as pathogenic (Secondary Dataset Disease). We supplemented this with 65 presumed benign variants from the BRAVO resource (Secondary Dataset Neutral) (**Figure 2**). All missense variants included in the primary and secondary analyses are shown in **Supplementary Table 2**.

### Descriptive analysis

When the distribution of the studied variants was mapped, presumed pathogenic changes tended to cluster around the two DNA-binding protein domains of PAX6: the Paired Domain (PD) and the HomeoDomain (HD). Conversely, presumed benign variants were more likely to affect residues outside these domains. VUS did not show a clear clustering pattern (**Figure 3**).

### Performance of computational tools

The predictive performance of ten tools was evaluated. When the performance metrics were calculated using the default threshold set by the tools’ developers, considerable variability was noted (**Table 1a**). Most tools exhibited high sensitivity (exceeding 88%) but had low specificity scores. SIFT4G and AlphaMissense

**Table 1.**
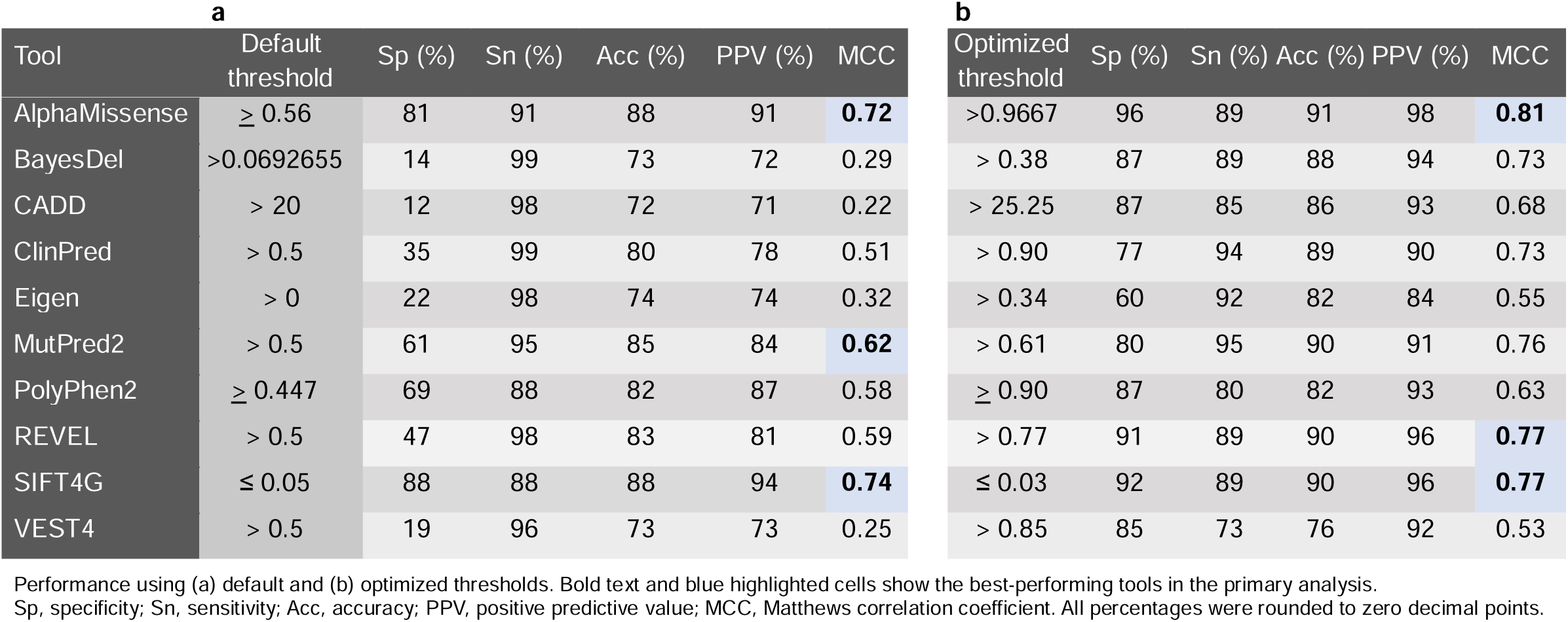
Performance of the computational tools assessed in this study (in tasks involving PAX6 missense variant evaluation)

| **a** |  |  |  |  |  |  | **b** |  |  |  |  |  |
| --- | --- | --- | --- | --- | --- | --- | --- | --- | --- | --- | --- | --- |
| Tool | Default threshold | Sp (%) | Sn (%) | Acc (%) | PPV (%) | MCC | Optimized threshold | Sp (%) | Sn (%) | Acc (%) | PPV (%) | MCC |
| AlphaMissense | $\geq 0.56$ | 81 | 91 | 88 | 91 | **0.72** | >0.9667 | 96 | 89 | 91 | 98 | **0.81** |
| BayesDel | >0.0692655 | 14 | 99 | 73 | 72 | 0.29 | > 0.38 | 87 | 89 | 88 | 94 | 0.73 |
| CADD | > 20 | 12 | 98 | 72 | 71 | 0.22 | > 25.25 | 87 | 85 | 86 | 93 | 0.68 |
| ClinPred | > 0.5 | 35 | 99 | 80 | 78 | 0.51 | > 0.90 | 77 | 94 | 89 | 90 | 0.73 |
| Eigen | > 0 | 22 | 98 | 74 | 74 | 0.32 | > 0.34 | 60 | 92 | 82 | 84 | 0.55 |
| MutPred2 | > 0.5 | 61 | 95 | 85 | 84 | **0.62** | > 0.61 | 80 | 95 | 90 | 91 | 0.76 |
| PolyPhen2 | $\geq 0.447$ | 69 | 88 | 82 | 87 | 0.58 | $\geq 0.90$ | 87 | 80 | 82 | 93 | 0.63 |
| REVEL | > 0.5 | 47 | 98 | 83 | 81 | 0.59 | > 0.77 | 91 | 89 | 90 | 96 | **0.77** |
| SIFT4G | $\leq 0.05$ | 88 | 88 | 88 | 94 | **0.74** | $\leq 0.03$ | 92 | 89 | 90 | 96 | **0.77** |
| VEST4 | > 0.5 | 19 | 96 | 73 | 73 | 0.25 | > 0.85 | 85 | 73 | 76 | 92 | 0.53 |

Performance using (a) default and (b) optimized thresholds. Bold text and blue highlighted cells show the best-performing tools in the primary analysis.
Sp, specificity; Sn, sensitivity; Acc, accuracy; PPV, positive predictive value; MCC, Matthews correlation coefficient. All percentages were rounded to zero decimal points.

achieved specificity scores of 88% and 81%, respectively. In contrast, other tools showed specificities below 70%, with CADD, BayesDel and VEST4 scoring the lowest at 12%, 14% and 19%, respectively. The other metrics, such as accuracy and PPV, ranged from 72% to 88% and 71% to 94%, respectively. The MCC scores ranged from 0.22 to 0.74, with the top-three tools attaining the highest scores being SIFT4G at 0.74, followed by AlphaMissense at 0.72 and MutPred2 at 0.62.

### Improving performance through threshold optimization

Aiming to obtain gene-specific thresholds tailored to *PAX6*, we performed ROC curve analysis and determined the value that achieved the maximum MCC score for each tool (see **Supplementary** Fig.1). The default thresholds were generally lower compared to the optimized thresholds (**Table 2b**), except for SIFT4G (which, unlike the other tools, assigns lower scores to variants with a higher likelihood of being predicted as pathogenic). Following threshold optimization, all the performance parameters of the tools showed improvement, with a notable increase in specificity scores. At the optimized threshold, AlphaMissense achieved the highest MCC score of 0.81, succeeded by SIFT4G and REVEL at 0.77 (**Table 2b**).

**Table 2.** Five-fold cross validation results showing the performance of the AlphaMissense tool (in tasks involving *PAX6* missense variant evaluation)

| Test | Sp (%) | Sn (%) | Acc (%) | PPV (%) | MCC |
| --- | --- | --- | --- | --- | --- |
| 1 | 93 | 94 | 93 | 97 | 0.85 |
| 2 | 86 | 97 | 93 | 94 | 0.84 |
| 3 | 93 | 84 | 87 | 96 | 0.74 |
| 4 | 93 | 82 | 85 | 96 | 0.70 |
| 5 | 100 | 91 | 94 | 100 | 0.87 |
| Average | 93.1 ± 5.1 | 89.5 ± 6.3 | 90.6 ± 4.0 | 96.7 ± 2.2 | 0.80 ± 0.1 |

Sp, specificity; Sn, sensitivity; Acc, Accuracy; PPV, positive predictive value; MCC, Matthews correlation coefficient. All percentages were rounded to zero decimal points.

### Performance of combination of tools

We assessed if the predictive performance could be further improved by combining multiple tools. A combination of the top-three tools (AlphaMissense, SIFT4G and REVEL) with optimized thresholds, demonstrated an MCC score of 0.78, with a sensitivity of 87% and accuracy of 90%. These results outperformed those obtained by combining the predictions of SIFT4G and AlphaMissense or REVEL and AlphaMissense but the MCC score was lower than the combination of SIFT4G and REVEL (**Supplementary Table 3**). Interestingly, the MCC score of AlphaMissense alone (following threshold optimization) was higher (0.81) than the MCC score of all combined approaches.

### Validation and further evaluation

To assess the reliability of the results of our primary analysis (concerning AlphaMissense), we conducted further studies using a five-fold cross-validation approach. The findings confirmed the robustness of AlphaMissense (with the threshold optimization) in predicting the effect of *PAX6* variants (**Table 2)**.

Further evaluation using a different set of variants (secondary dataset) confirmed (i) that AlphaMissense and SIFT4G are among the higher-ranking tools; and (ii) that gene-specific thresholds lead to enhanced predictive performance (**Table 3**). It is worth noting that, except for sensitivity, the values in the secondary analysis were lower than those obtained in the primary analysis. This difference is likely to be influenced by the varying proportion of presumed benign and presumed pathogenic variants between the corresponding primary and secondary datasets.

**Table 3.** Performance of the computational tools assessed in this study (in tasks involving PAX6 missense variant evaluation): secondary analysis.

| Tool | Threshold | Sp (%) | Sn (%) | Acc (%) | PPV (%) | MCC |
| --- | --- | --- | --- | --- | --- | --- |
| AlphaMissense | Default thresholds | 65 | 100 | 69 | 30 | 0.44 |
| BayesDel |  | 6 | 100 | 19 | 14 | 0.09 |
| CADD |  | 3 | 100 | 16 | 14 | 0.06 |
| ClinPred |  | 12 | 100 | 24 | 15 | 0.14 |
| Eigen |  | 15 | 100 | 26 | 14 | 0.14 |
| MutPred2 |  | 34 | 100 | 43 | 19 | 0.25 |
| PolyPhen2 |  | 65 | 100 | 69 | 30 | 0.44 |
| REVEL |  | 37 | 100 | 45 | 20 | 0.27 |
| SIFT4G |  | 79 | 90 | 80 | 39 | 0.50 |
| VEST4 |  | 20 | 100 | 31 | 16 | 0.18 |
| AlphaMissense | Optimized thresholds | 88 | 90 | 88 | 53 | **0.63** |
| REVEL |  | 82 | 100 | 84 | 46 | 0.61 |
| SIFT4G |  | 83 | 90 | 84 | 45 | 0.56 |
| Combination (AlphaMissense + SIFT4G + REVEL) |  | 85 | 90 | 85 | 47 | 0.58 |

Bold text and blue highlighted cell shows the best-performing tool in the secondary analysis.
Sp, specificity; Sn, sensitivity; Acc, accuracy; PPV, positive predictive value; MCC, Matthews correlation coefficient. All percentages were rounded to zero decimal points.

Lastly, a set of seven VUS from our local database were analysed. Among these variants, six were consistently classified as pathogenic by all the ten tools investigated. However, one variant, *PAX6* c.926T>G, p.(Phe309Cys), showed discordant predictions (see **Supplementary Table 2b**). AlphaMissense and SIFT4G labelled this variant as predicted benign (with scores of 0.1654 and 0.16, respectively), while the other six variants were classified as predicted pathogenic. Notably, *PAX6* c.926T>G, p.(Phe309Cys), affects a residue in the C-terminal region, whereas the other six variants alter residues in one of the *PAX6* DNA-binding domains (PD or HD).

## DISCUSSION

We assessed the performance of ten commonly used variant prediction tools in the context of missense variants in a highly-conserved gene, *PAX6*. Using default settings, most tools were able to make reliable predictions in relation to pathogenic variants. However, their ability to correctly predict benign variants was limited (*i.e.*, there was high sensitivity but low specificity). These results are consistent with those from previous studies conducted on a genome-wide or an individual gene level.^13,37,38^ By generating optimized, gene-specific thresholds for each tool, it was possible to achieve improved performance compared to conventional approaches.

When default thresholds were used, SIFT4G, AlphaMissense and MutPred2 were found to be the top-ranking algorithms (*i.e.*, had the highest MCC scores). Following threshold optimization, AlphaMissense emerged as the best performing tool with the highest MCC score, followed by SIFT4G and REVEL, while MutPred2 shifted to the fourth position. AlphaMissense uses a deep learning model that builds on the protein structure prediction tool AlphaFold2.^24^ SIFT4G evaluates the impact of amino acid substitutions based on evolutionary conservation and sequence homology, aligning well with the highly-conserved nature of the *PAX6* gene.^25,39^ MutPred2 also incorporates a conservation-based approach along with other features. It is noted that MutPred2 was previously found to have good performance in prediction tasks involving variants in *PITX2*, a paired-like homeodomain transcription factor that is also expressed in the developing eye.^40^ REVEL emerged as the best meta-predictor in the context of *PAX6*; this was unsurprising as its superior performance over other ensemble tools has previously been demonstrated.^38,41,42,43^

Our findings support the use of gene-specific thresholds, as opposed to relying on default settings.^44^ Even REVEL, one of the highest performing tools, had a specificity of 47% (misclassifying 39 out of 74 presumed benign missense variants) with the default threshold. This issue arises due to the training process of the tools, where variants from multiple genes are used. This default approach allows for the possibility of underfitting, where crucial details necessary to capture the characteristics of an individual gene are overlooked. It is noted that, upon applying optimized thresholds, all tools demonstrated substantial improvement, particularly in specificity (**Table 2**). This observation is consistent with the findings of other studies looking at different genes.^11,13^

We attempted to combine the predictions of the top-three performing tools (following threshold optimization) using the majority-rule method. The results demonstrated good performance, with most of the parameters surpassing 84% and the MCC ranging from 0.76 to 0.79 (**Supplementary Table 3**). However, the use of AlphaMissense alone outperformed this approach (**Table 1b**). The high performance of this tool was confirmed through a 5-fold cross-validation experiment and in the secondary dataset (**Table 3**). To a degree, our findings contradict the observations of similar studies. For instance, Leong *et al.* found that the best performance for predicting *KCNQ1* variant pathogenicity was achieved by considering three out of the five tools that were examined.^12^ Likewise, Tang *et al.* reported achieving optimal performance in the context of *SCN1A* variants when combining the three best-performing tools.^11^ Conversely, our findings align with those of a study by Gunning *et al.* which supported the adoption of a single tool instead of using a consensus-based approach.^41^

Using AlphaMissense to evaluate seven *PAX6* missense variants that have been previously classified as VUS resolved some of the discordance for one change, c.926T>G, p.(Phe309Cys), by suggesting that it does not have an effect on molecular function. This variant, unlike most *PAX6* pathogenic missense changes, affects a residue outside the DNA-binding domains.^45^ This result could potentially be attributed to AlphaMissense’s ability to pinpoint functionally crucial sites (instead of simply evaluating the overall evolutionary conservation of a protein).^24^ It is noted that a few recent studies have shown that AlphaMissense can reliably classify subsets of variants that are known to affect molecular function.^46–48^

The present study has several limitations, including the availability of a relatively small number of presumed pathogenic variants due to the rarity of *PAX6*-related disease. Additionally, we were unable to exclude the possibility that some of the studied genetic variants may have been utilized for training some of the evaluated tools. Future studies could explore the performance of a wider range of computational approaches, including tools considering the 3D-structure of the protein and algorithms using advanced artificial neural network approaches.

It is highlighted that variant pathogenicity predictors constitute one of the many pieces of evidence that can be used to evaluate the effect of genetic alterations, and that it is crucial to consider other factors (including segregation analysis, population frequency and the outcomes of functional assays).^49^ Additionally, refinement of the ACMG/AMP sequence variant guidelines (and utilization of Bayesian approaches) is expected to provide an enhanced framework that would help generate robust estimates by improving how different lines of evidence are combined.

## CONCLUSION

In summary, this study offers insights into how computational prediction tools can be optimally used for the task of *PAX6* missense variant evaluation. The best-performing approach, which involves using a *PAX6*-specific threshold for AlphaMissense, can be utilized in different contexts and has the potential to enhance variant interpretation, ultimately leading to more precise and timely diagnoses for individuals with *PAX6*-related disorders.

## Data Availability

All data produced in the present work are included in the manuscript and the supplementary files

## SUPPLEMENTARY INFORMATION

Supplementary Figure 1

Supplementary Tables 1-3

## ACKNOWLEDGEMENTS & FUNDING

We acknowledge the following sources of funding: the Wellcome Trust (224643/Z/21/Z, Clinical Research Career Development Fellowship to P.I.S.; 200990/Z/16/Z, Transforming Genetic Medicine Initiative to G.C.B.); the UK National Institute for Health Research (NIHR) Clinical Lecturer Programme (CL-2017-06- 001652 to P.I.S.); Retina UK and Fight for Sight (GR586, RP Genome Project - UK Inherited Retinal Disease Consortium to G.C.B.); and the Indonesia Endowment Fund for Education (Lembaga Pengelola Dana Pendidikan (LPDP) scholarship to N.S.A.). This research was co-funded by the NIHR Manchester Biomedical Research Centre (NIHR203308). The views expressed are those of the author(s) and not necessarily those of the NIHR or the Department of Health and Social Care.

## MAIN WEB RESOURCES

- Genome Aggregation Database version 2.1.1 (v2) and version 3.1.1 (v3) https://gnomad.broadinstitute.org/, accessed in February 2023
- Leiden Open Variation Database version 2.0 and 3.0 https://www.lovd.nl/, accessed in February 2023
- Human Genetic Mutation Database https://www.hgmd.cf.ac.uk/, accessed in February 2023
- ClinVar https://www.ncbi.nlm.nih.gov/clinvar/, accessed in February 2023
- BRAVO Powered by TOPMed Freeze 8 on GRCh38 https://bravo.sph.umich.edu/freeze8/hg38/, accessed in April 2023
- cBioPortal https://www.cbioportal.org/, accessed in May 2023
- dbNSFP http://database.liulab.science/dbNSFP, accessed in March 2023

## ABBREVIATIONS

Acc: accuracy
ACMG/AMP: the American College of Medical Genetics and Genomics and the Association for Molecular Pathology
AUC: area under the ROC curve
dbNSFP: database of non-synonymous functional prediction
DM: disease-causing mutation
DM?: likely disease-causing mutation with questionable pathogenicity
gnomAD: Genome Aggregation Database
GRCh38: genome reference consortium human build 38
HD: homeodomain
HGMD: Human Gene Mutation Database
HGNC: HUGO Gene Nomenclature Committee
LOVD: Leiden Open Variation Database
MCC: Matthews correlation coefficient
MCGM: Manchester Centre for Genomic Medicine
OMIM: Online Mendelian Inheritance in Man
PD: paired domain
PAX 6: paired box 6
PPV: positive predictive value
ROC: receiver operating characteristic
Sn: sensitivity
Sp: specificity
VUS: variant of uncertain significance

## REFERENCES

1. Walther C, Gruss P. Pax-6, a murine paired box gene, is expressed in the developing CNS. Development. 1991;113(4):1435–49. DOI: 10.1242/dev.113.4.1435

2. Mishra R, Gorlov IP, Chao LY, Singh S, Saunders GF. PAX6, Paired Domain Influences Sequence Recognition by the Homeodomain*. J Biol Chem. 2002;277(51):49488–94. DOI: 10.1074/jbc.M206478200

3. Moosajee M, Hingorani M, Moore AT. PAX6-Related Aniridia. In: Adam MP, Mirzaa GM, Pagon RA, Wallace SE, Bean LJ, Gripp KW, et al., editors. GeneReviews® [Internet]. Seattle (WA): University of Washington, Seattle; 1993 [cited 2023 Jul 7]. Available from: http://www.ncbi.nlm.nih.gov/books/NBK1360/

4. Tzoulaki I, White IM, Hanson IM. PAX6 mutations: genotype-phenotype correlations. BMC Genet. 2005;6(1):27. DOI: 10.1186/1471-2156-6-27

5. Hanson I, Churchill A, Love J, Axton R, Moore T, Clarke M, et al. Missense mutations in the most ancient residues of the PAX6 paired domain underlie a spectrum of human congenital eye malformations. Hum Mol Genet. 1999;8(2):165–72. DOI: 10.1093/hmg/8.2.165

6. Williamson KA, Hall HN, Owen LJ, Livesey BJ, Hanson IM, Adams G, et al. Recurrent heterozygous PAX6 missense variants cause severe bilateral microphthalmia via predictable effects on DNA–protein interaction. Genet Med. 2020;22(3):598–609. DOI: 10.1038/s41436-019-0685-9

7. Cross E, Duncan-Flavell PJ, Howarth RJ, Crooks RO, Thomas NS, Bunyan DJ. Screening of a large PAX6 cohort identified many novel variants and emphasises the importance of the paired and homeobox domains. Eur J Med Genet. 2020;63(7):103940. DOI: 10.1016/j.ejmg.2020.103940

8. Richards S, Aziz N, Bale S, Bick D, Das S, Gastier-Foster J, et al. Standards and guidelines for the interpretation of sequence variants: a joint consensus recommendation of the American College of Medical Genetics and Genomics and the Association for Molecular Pathology. Genet Med. 2015;17(5):405–24. DOI: 10.1038/gim.2015.30

9. Liu Y, Yeung WSB, Chiu PCN, Cao D. Computational approaches for predicting variant impact: An overview from resources, principles to applications. Front Genet. 2022;13:981005. DOI: 10.3389/fgene.2022.981005

10. Tamana S, Xenophontos M, Minaidou A, Stephanou C, Harteveld CL, Bento C, et al. Evaluation of in silico predictors on short nucleotide variants in HBA1, HBA2, and HBB associated with haemoglobinopathies. eLife. 2022;11:e79713. DOI: 10.7554/eLife.79713

11. Tang B, Li B, Gao LD, He N, Liu XR, Long YS, et al. Optimization of in silico tools for predicting genetic variants: individualizing for genes with molecular sub-regional stratification. Brief Bioinform. 2020;21(5):1776–86. DOI: 10.1093/bib/bbz115

12. Leong IU, Stuckey A, Lai D, Skinner JR, Love DR. Assessment of the predictive accuracy of five in silico prediction tools, alone or in combination, and two metaservers to classify long QT syndrome gene mutations. BMC Med Genet. 2015;16(1):34. DOI: 10.1186/s12881-015-0176-z

13. Sallah SR, Ellingford JM, Sergouniotis PI, Ramsden SC, Lench N, Lovell SC, et al. Improving the clinical interpretation of missense variants in X linked genes using structural analysis. J Med Genet. 2022;59(4):385–92. DOI: 10.1136/jmedgenet-2020-107404

14. Amendola LM, Jarvik GP, Leo MC, McLaughlin HM, Akkari Y, Amaral MD, et al. Performance of ACMG-AMP Variant-Interpretation Guidelines among Nine Laboratories in the Clinical Sequencing Exploratory Research Consortium. Am J Hum Genet. 2016;99(1):247. DOI: 10.1016/j.ajhg.2016.03.024

15. The Critical Assessment of Genome Interpretation Consortium. CAGI, the Critical Assessment of Genome Interpretation, establishes progress and prospects for computational genetic variant interpretation methods [Internet]. arXiv; 2022 [cited 2023 Jul 27]. Available from: http://arxiv.org/abs/2205.05897

16. Gudmundsson S, Singer-Berk M, Watts NA, Phu W, Goodrich JK, Solomonson M, et al. Variant interpretation using population databases: Lessons from gnomAD. Hum Mutat. 2022;43(8):1012–30. DOI: 10.1002/humu.24309

17. Fokkema IFAC, Taschner PEM, Schaafsma GCP, Celli J, Laros JFJ, den Dunnen JT. LOVD v.2.0: the next generation in gene variant databases. Hum Mutat. 2011;32(5):557–63. DOI: 10.1002/humu.21438

18. Fokkema IFAC, Kroon M, López Hernández JA, Asscheman D, Lugtenburg I, Hoogenboom J, et al. The LOVD3 platform: efficient genome-wide sharing of genetic variants. Eur J Hum Genet. 2021;29(12):1796–803. DOI: 10.1038/s41431-021-00959-x

19. Stenson PD, Mort M, Ball EV, Chapman M, Evans K, Azevedo L, et al. The Human Gene Mutation Database (HGMD®): optimizing its use in a clinical diagnostic or research setting. Hum Genet. 2020;139(10):1197–207. DOI: 10.1007/s00439-020-02199-3

20. Landrum MJ, Lee JM, Benson M, Brown GR, Chao C, Chitipiralla S, et al. ClinVar: improving access to variant interpretations and supporting evidence. Nucleic Acids Res. 2018;46(D1):D1062–7. DOI: 10.1093/nar/gkx1153

21. Taliun D, Harris DN, Kessler MD, Carlson J, Szpiech ZA, Torres R, et al. Sequencing of 53,831 diverse genomes from the NHLBI TOPMed Program. Nature. 2021;590(7845):290–9. DOI: 10.1038/s41586-021-03205-y

22. The UniProt Consortium. UniProt: the Universal Protein Knowledgebase in 2023. Nucleic Acids Res. 2023;51(D1):D523–31. DOI: 10.1093/nar/gkac1052

23. Vohra S, Biggin PC. Mutationmapper: A Tool to Aid the Mapping of Protein Mutation Data. PLoS One. 2013;8(8):e71711. DOI: 10.1371/journal.pone.0071711

24. Cheng J, Novati G, Pan J, Bycroft C, Žemgulytė A, Applebaum T, et al. Accurate proteome-wide missense variant effect prediction with AlphaMissense. Science. 2023;381(6664):eadg7492. DOI: 10.1126/science.adg7492

25. Vaser R, Adusumalli S, Leng SN, Sikic M, Ng PC. SIFT missense predictions for genomes. Nat Protoc. 2016;11(1):1–9. DOI: 10.1038/nprot.2015.123

26. Adzhubei I, Jordan DM, Sunyaev SR. Predicting Functional Effect of Human Missense Mutations Using PolyPhen-2. Curr Protoc Hum Genet. 2013;76(1):7.20.1–7.20.41. DOI: 10.1002/0471142905.hg0720s76

27. Carter H, Douville C, Stenson PD, Cooper DN, Karchin R. Identifying Mendelian disease genes with the Variant Effect Scoring Tool. BMC Genom. 2013;14(Suppl 3):S3. DOI: 10.1186/1471-2164-14-S3-S3

28. Ioannidis NM, Rothstein JH, Pejaver V, Middha S, McDonnell SK, Baheti S, et al. REVEL: An Ensemble Method for Predicting the Pathogenicity of Rare Missense Variants. Am J Hum Genet. 2016;99(4):877–85. DOI: 10.1016/j.ajhg.2016.08.016

29. Kircher M, Witten DM, Jain P, O’Roak BJ, Cooper GM, Shendure J. A general framework for estimating the relative pathogenicity of human genetic variants. Nat Genet. 2014;46(3):310–5. DOI: 10.1038/ng.2892

30. Pejaver V, Urresti J, Lugo-Martinez J, Pagel KA, Lin GN, Nam HJ, et al. Inferring the molecular and phenotypic impact of amino acid variants with MutPred2. Nat Commun. 2020;11(1):5918. DOI: 10.1038/s41467-020-19669-x

31. Feng BJ. PERCH: A Unified Framework for Disease Gene Prioritization. Hum Mutat. 2017;38(3):243–51. DOI: 10.1002/humu.23158

32. Alirezaie N, Kernohan KD, Hartley T, Majewski J, Hocking TD. ClinPred: Prediction Tool to Identify Disease-Relevant Nonsynonymous Single-Nucleotide Variants. Am J Hum Genet. 2018;103(4):474–83. DOI: 10.1016/j.ajhg.2018.08.005

33. Ionita-Laza I, McCallum K, Xu B, Buxbaum JD. A spectral approach integrating functional genomic annotations for coding and noncoding variants. Nat Genet. 2016;48(2):214–20. DOI: 10.1038/ng.3477

34. Liu X, Li C, Mou C, Dong Y, Tu Y. dbNSFP v4: a comprehensive database of transcript-specific functional predictions and annotations for human nonsynonymous and splice-site SNVs. Genome Med. 2020;12(1):103. DOI: 10.1186/s13073-020-00803-9

35. Niroula A, Vihinen M. Variation Interpretation Predictors: Principles, Types, Performance, and Choice. Hum Mutat. 2016;37(6):579–97. DOI: 10.1002/humu.22987

36. IBM Corp. IBM SPSS Statistics for Windows. Armonk, NY: IBM Corp; 2021.

37. Ernst C, Hahnen E, Engel C, Nothnagel M, Weber J, Schmutzler RK, et al. Performance of in silico prediction tools for the classification of rare BRCA1/2 missense variants in clinical diagnostics. BMC Med Genomics. 2018;11(1):35. DOI: 10.1186/s12920-018-0353-y

38. Li J, Zhao T, Zhang Y, Zhang K, Shi L, Chen Y, et al. Performance evaluation of pathogenicity-computation methods for missense variants. Nucleic Acids Res. 2018;46(15):7793–804. DOI: 10.1093/nar/gky678

39. Ng PC, Henikoff S. SIFT: predicting amino acid changes that affect protein function. Nucleic Acids Res. 2003;31(13):3812–4. DOI: 10.1093/nar/gkg509

40. Seifi M, Walter MA. Accurate prediction of functional, structural, and stability changes in PITX2 mutations using in silico bioinformatics algorithms. Cai T, editor. PLoS One. 2018;13(4):e0195971. DOI: 10.1371/journal.pone.0195971

41. Gunning AC, Fryer V, Fasham J, Crosby AH, Ellard S, Baple EL, et al. Assessing performance of pathogenicity predictors using clinically relevant variant datasets. J Med Genet. 2021;58(8):547–55. DOI: 10.1136/jmedgenet-2020-107003

42. Tian Y, Pesaran T, Chamberlin A, Fenwick RB, Li S, Gau CL, et al. REVEL and BayesDel outperform other in silico meta-predictors for clinical variant classification. Sci Rep. 2019;9(1):12752. DOI: 10.1038/s41598-019-49224-8

43. Hopkins JJ, Wakeling MN, Johnson MB, Flanagan SE, Laver TW. REVEL is better at predicting pathogenicity of loss-of-function than gain-of-function variants [Internet]. medRxiv; 2023 [cited 2023 Jul 26]. p. 2023.06.06.23290963. Available from: https://www.medrxiv.org/content/10.1101/2023.06.06.23290963v1

44. Pejaver V, Byrne AB, Feng BJ, Pagel KA, Mooney SD, Karchin R, et al. Calibration of computational tools for missense variant pathogenicity classification and ClinGen recommendations for PP3/BP4 criteria. Am J Hum Genet. 2022;109(12):2163–77. DOI: 10.1016/j.ajhg.2022.10.013

45. Laddach A, Ng JCF, Fraternali F. Pathogenic missense protein variants affect different functional pathways and proteomic features than healthy population variants. PLoS Biol. 2021;19(4):e3001207. DOI: 10.1371/journal.pbio.3001207

46. Tordai H, Torres O, Csepi M, Padányi R, Lukács GL, Hegedűs T. Lightway access to AlphaMissense data that demonstrates a balanced performance of this missense mutation predictor [Internet]. Bioinformatics; 2023 Nov [cited 2023 Nov 20]. Available from: http://biorxiv.org/lookup/doi/10.1101/2023.10.30.564807

47. Staklinski SJ, Scheben A, Siepel A, Kilberg MS. Utility of AlphaMissense predictions in Asparagine Synthetase deficiency variant classification [Internet]. Genetics; 2023 Nov [cited 2023 Nov 20]. Available from: http://biorxiv.org/lookup/doi/10.1101/2023.10.30.564808

48. Ljungdahl A, Kohani S, Page NF, Wells ES, Wigdor EM, Dong S, et al. AlphaMissense is better correlated with functional assays of missense impact than earlier prediction algorithms [Internet]. bioRxiv; 2023 [cited 2023 Dec 3]. p. 2023.10.24.562294. Available from: https://www.biorxiv.org/content/10.1101/2023.10.24.562294v1

49. Garcia FADO, Andrade ESD, Palmero EI. Insights on variant analysis in silico tools for pathogenicity prediction. Front Genet. 2022;13:1010327. DOI: 10.3389/fgene.2022.1010327

