## Supplementary Figure 1 for "Using computational approaches to enhance the interpretation of missense variants in the *PAX6* gene"

**Supplementary Figure 1**. Receiver operating characteristic (ROC) curves for the computational tools assessed in this study (in tasks involving *PAX6* missense variant evaluation).

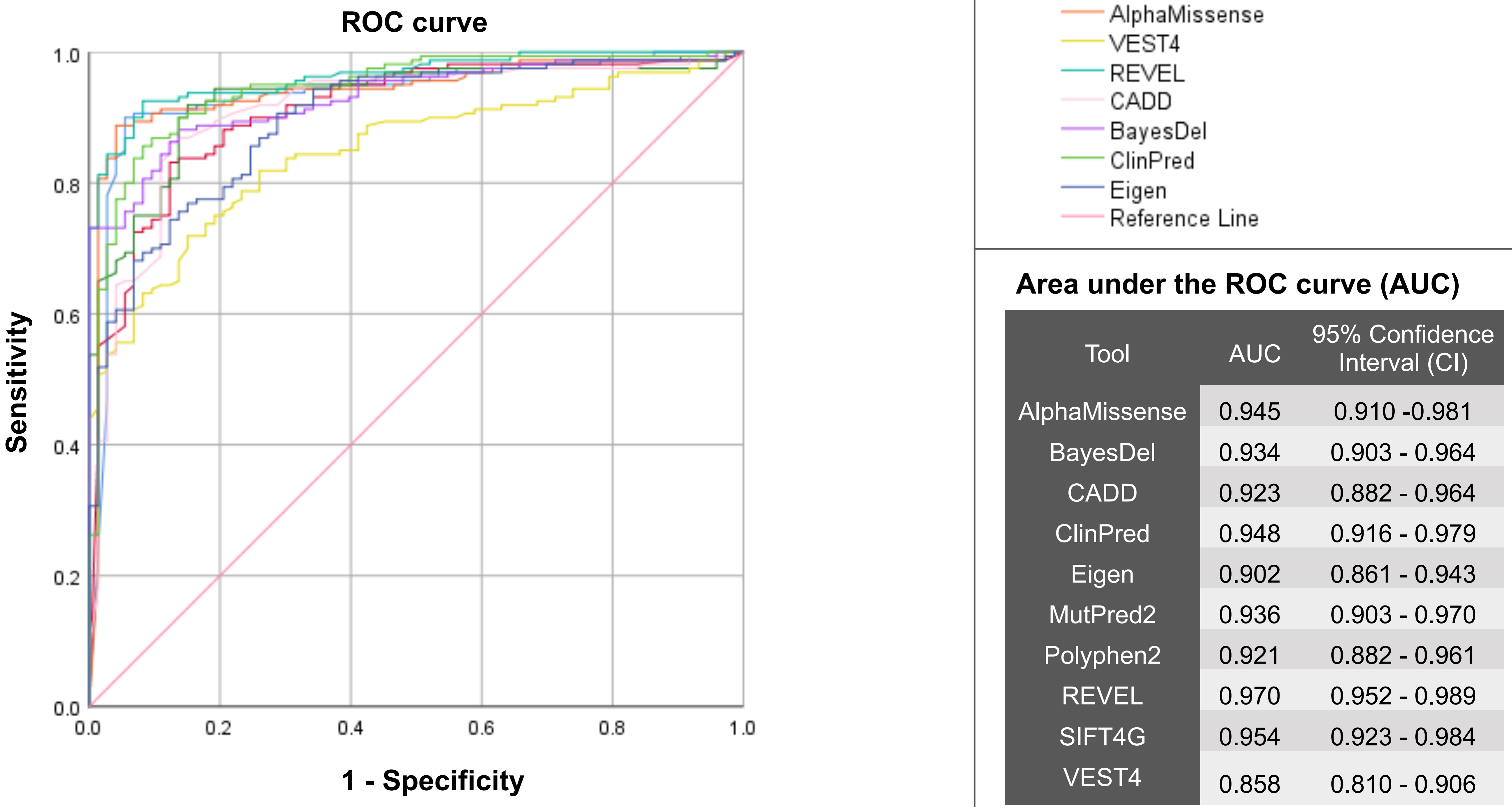


**Area under the ROC curve (AUC)**

| Tool | AUC | 95% Confidence Interval (CI) |
|---|---|---|
| AlphaMissense | 0.945 | 0.910 -0.981 |
| BayesDel | 0.934 | 0.903 - 0.964 |
| CADD | 0.923 | 0.882 - 0.964 |
| ClinPred | 0.948 | 0.916 - 0.979 |
| Eigen | 0.902 | 0.861 - 0.943 |
| MutPred2 | 0.936 | 0.903 - 0.970 |
| Polyphen2 | 0.921 | 0.882 - 0.961 |
| REVEL | 0.970 | 0.952 - 0.989 |
| SIFT4G | 0.954 | 0.923 - 0.984 |
| VEST4 | 0.858 | 0.810 - 0.906 |
