## Supplementary Table 1 for "Using computational approaches to enhance the interpretation of missense variants in the *PAX6* gene"

**Supplementary Table 1.** Computational tools assessed in this study

| Approach | Tool | Description | Output |
|---|---|---|---|
| **Combination of structural context and evolutionary conservation** | AlphaMissense | • Training on weak labels from population frequency data, which avoid the bias of using human annotations.<br>• An unsupervised protein language model, learning the amino acid distributions based on sequence contexts.<br>• Considering structural contexts derived from AlphaFold, incorporating protein three-dimensional information. | ▪ score range: 0-1<br>▪ benign: 0.00-0.33<br>▪ ambiguous: 0.34-0.56<br>▪ pathogenic: 0.57-1 |
| **Unsupervised machine learning** | Eigen | • Utilises a support vector machine and incorporates evolutionary parameters, ENCODE summaries, and population frequencies from the 1000 Genomes project, with an unsupervised approach.<br>• Generates Eigen PC scores for non-coding variants.<br>• Both Eigen and Eigen-PC exhibit similar performance for coding variants, with Eigen demonstrating slightly superior performance. | ▪ score range: from below 0 to over 1; threshold set at 0.<br>▪ higher scores indicate a higher likelihood of a pathogenic prediction |
| **Sequence / structure** | MutPred2 | • Compared to the previous version, MutPred, the tool forecasts alterations in particular protein properties resulting from mutations, such as catalytic activity, stability, and protein-protein interactions.<br>• Utilises a neural network ensemble to create a classification function that generates more balanced scores than the random forest model. The tool also aims to uncover the molecular factors responsible for detrimental amino acid substitutions. | ▪ score range: 0-1<br>▪ benign: < 0.5<br>▪ damaging: ≥ 0.5 |
| | PolyPhen2 | • Utilises a naïve Bayesian approach and incorporates information from annotated UniProt entries to ascertain if a missense mutation resides in a protein region that is structurally significant or functionally crucial.<br>• Examines multiple features, including sequence composition, active binding sites, structure-based properties, and the effect of the variant on factors like accessible area, hydrophobicity, chemical-electrostatic interactions, secondary structure conformation, solvent accessible surface area, and Phi-Psi dihedral angles. | ▪ score range: 0-1<br>▪ probably damaging: 0.909-1<br>▪ possibly damaging: 0.447-0.908<br>▪ benign: 0-0.446 |
| **Homology / conservation** | SIFT4G | • An enhanced iteration and a faster version of SIFT (Sorts Intolerant from Tolerant) that employs sequence homology derived from multiple sequence alignments.<br>• Operates under the assumption that functionally significant missense variants typically occur at evolutionarily conserved sites, while most single nucleotide polymorphisms are considered neutral. | ▪ score range: 0-1<br>▪ damaging: ≤ 0.05<br>▪ tolerated: > 0.5 |

(continued)

(continued)

| Approach | Tool | Description | Output |
|---|---|---|---|
| **Supervised machine learning** | BayesDel | • Combines annotation scores using the naïve Bayesian approach derived from PolyPhen2, SIFT, FATHMM, LRT, Mutation Taster, Mutation Assessor, PhyloP, GERP++, Siphy. Conservation measures and minor allele frequencies across different populations (from ExAC (Exome Aggregation Consortium)) are included.<br>• The integration of these scores results in a combined deleteriousness score, which is defined as a weighted product of likelihood ratios. | ▪ scores range from below 0 to over 1; threshold set at 0.0692655.<br>▪ higher scores indicate a higher likelihood of a pathogenic prediction |
| | CADD (Combined Annotation-Dependent Depletion) | • Incorporates diverse sources such as conservation matrices (GERP, PhastCons, PhyloP), functional annotations, protein-level scores (Grantham, SIFT, PolyPhen2), and data from genomic studies including gene expression values, acetylation, methylation, nucleosome occupancy, chromatin status, transcription factors, 1,000 Genomes, and Exome Sequencing Project.<br>• Employs a support vector machine to evaluate single nucleotide variants or small insertions/ deletions | ▪ Phred score range 1-99; threshold set at 20.<br>▪ higher scores indicate a higher likelihood of a pathogenic prediction |
| | ClinPred | • Utilises a combination of two machine learning algorithms, random forest and gradient-boosted decision tree models.<br>• Integrates scores from 16 tools, including SIFT, PolyPhen-2 HDIV, PolyPhen-2 HVAR, LRT, MutationAssessor, PROVEAN, CADD, GERP, DANN, PhastCons, fitCons, PhyloP, and SiPhy.<br>• Incorporates normal population allele frequency data from gnomAD. | ▪ score range: 0-1<br>▪ benign: < 0.5<br>▪ damaging: ≥ 0.5 |
| | REVEL (Rare Exome Variant Ensemble Learning) | • A meta-predictor that integrates 18 pathogenic prediction scores from 13 tools (GERP++, SIFT, PolyPhen-2, MutationTaster, Mutation Assessor, FATHMM, LRT, PhyloP, PhastCons, SIPHY, MutPred, PROVEAN, and VEST) alongside eight sequence conservation scores and ten functional scores.<br>• Users have the flexibility to customise the pathogenicity threshold based on their preferences (0.5 as default, 0,7 for more stringent cut-off). | ▪ score range: 0-1<br>▪ benign: < 0.5<br>▪ damaging: ≥ 0.5 |
| | VEST4 (Variant Effect Scoring Tool 4.0) | • Utilises the random forest method to predict the impact of gene malfunction and its associated mutations.<br>• Takes into account the contextual aspect of diverse variations between disease-specific and commonly occurring variants.<br>• Not an ensemble approach unlike the other tools in this algorithm; instead, the tool uses 86 characteristics as its input. | ▪ score range: 0-1<br>▪ benign: < 0.5<br>▪ damaging: ≥ 0.5 |
