## Supplementary Table 3 for "Using computational approaches to enhance the interpretation of missense variants in the *PAX6* gene"

**Supplementary Table 3.** Performance of the computational tool combinations assessed in this study (in tasks involving *PAX6* missense variant evaluation).

| Combination | Sp (%) | Sn (%) | Acc (%) | PPV (%) | MCC |
|---|---|---|---|---|---|
| AlphaMissense + REVEL | 96 | 85 | 88 | 98 | 0.76 |
| AlphaMissense + SIFT4G | 96 | 84 | 88 | 98 | 0.76 |
| REVEL + SIFT4G | 97 | 86 | 90 | 99 | 0.79 |
| AlphaMissense + REVEL + SIFT4G | 96 | 87 | 90 | 98 | 0.78 |

A genetic variant was considered pathogenic if the scores of ≥ 2 tools surpassed the corresponding optimized thresholds. Sp, specificity; Sn, sensitivity; Acc, Accuracy; PPV, positive predictive value; MCC, Matthews correlation coefficient.  All percentages were rounded to zero decimal points.
